# Changing parental perceptions to childhood immunisations during the Covid-19 pandemic in the UK: evidence from three cross-sectional surveys between 2020 and 2022

**DOI:** 10.1101/2024.11.29.24318181

**Authors:** Rosie Thistlethwayte, Alexandre de Figueiredo

## Abstract

**OBJECTIVE:** Childhood immunisation rates in the UK have recently fallen to their lowest level in 14 years. There is currently a lack of temporal evidence on parental attitudes to childhood immunisations and how they have evolved in response to the Covid-19 pandemic, limiting our ability to assess the impact of the pandemic on population-level attitudes to non-Covid vaccines. This study aims to assess trends in parental confidence in childhood immunisations between 2020 and 2022 at varying spatial scales in the UK, while also identifying the socio-demographic factors associated with vaccine perceptions and how these have shifted over time.

**DESIGN:** Three cross-sectional surveys in 2020, 2021, and 2022.

**SETTING:** United Kingdom.

**PARTICIPANTS:** 14,720 adults responsible for decisions surrounding the vaccination of children.

**MAIN OUTCOME MEASURES:** The percentage of parents indicating past or future refusal for the MMR, HPV, and influenza vaccines for their child in 2020, 2021, and 2022 as well as Covid-19 vaccine refusal for their child in 2022. A combined metric (refusal) is created to measure parental refusal for any childhood vaccine. Associations between these primary outcome measures and socio-demographic variables are investigated via multiple logistic regression, with effects reported via odds ratios. Additionally, the extent to which parental and caregiver perceptions in giving children immunisations since the start of the pandemic is examined using data from 2022.

**RESULTS:** Across the UK overall refusal decreased from 16.2% in 2020 to 14.0% in 2021 (*p*<0.001) before increasing to 20.8% in 2022 (*p*<0.001 compared to 2020). This loss was driven by relatively high rates of parental refusal of the Covid-19 vaccine for their children in 2022, rather than perceptions towards other childhood vaccines. A marked negative change in perceptions towards giving childhood vaccines is found among parents who had not themselves received at least three doses of a Covid-19 vaccine, signalling a strong spillover effect of Covid-19 vaccine hesitancy onto routine childhood vaccines. Many parental socio-demographic factors were found to be informative of vaccine refusal, with younger age groups, individuals living in Greater London, Hindus, and Muslims exhibiting higher rates of refusal. Interestingly, however, 18— 34-year-olds, Hindus, Muslims, and Black / Black British respondents report becoming more positive towards giving about giving their children vaccines in 2022 compared to the start of the pandemic.

**CONCLUSIONS:** The available evidence suggests that parental refusal of routine childhood immunisations has decreased between 2020 and 2022 and remains low across the UK. Encouragingly, many socio-demographic groups with historically low confidence in childhood immunisations appear to be more positive about giving their children vaccines in 2022 compared to the beginning of the pandemic. While these findings are cautiously optimistic, there is still a mismatch between these reported increases in vaccine confidence and uptake rates of routine immunisations across the UK. Parents who did not receive at least three doses of a Covid-19 vaccine feel much less positive about giving their children vaccines since the start of the pandemic compared to those who did receive at least three doses. This group represents an important cohort for targeted outreach and tailored interventions to address lingering concerns and support vaccine uptake.

## Introduction

Since the start of the Covid-19 pandemic, declining childhood vaccination rates^1,2^ have raised concerns about the extent to which the reduced uptake is due to falling vaccine confidence or pandemic-induced practical barriers. Data from the UK Health Security Agency (UKHSA) show that routine childhood vaccine uptake in the UK remains below pre-pandemic levels,^3^ with measles coverage particularly alarming, as it continues to fall short of the WHO’s 95% target for elimination.^4,5^ The UKHSA’s June 2024 vaccine coverage update^6^ highlights further declines in vaccines administered before 24 months of age, with the measles, mumps, and rubella (MMR) vaccine coverage lagging behind other routine immunisations. MMR coverage in England has steadily decreased over the past decade, with second-dose MMR (MMR2) coverage dropping from 88.6% in 2014-15^7^ to just under 85% by late 2023.^6^

Of notable concern in the UK is the significant geographic variation in vaccine uptake, especially in regions like London, North West England, and the East Midlands^1^. In Hackney, MMR2 coverage is as low as 60.4%, and only five of 33 local authorities in London have achieved coverage above 80%^1^. Disruptions to healthcare services^8^, pandemic restrictions,^9^ and the reallocation of resources^10^ have been cited as reasons for declining routine immunisation rates during the pandemic. During the first national lockdown, a survey of 1,252 parents by Bell et al. found that fear of catching Covid-19 and confusion over the availability of vaccination services were key factors driving low vaccine uptake.^11^ Although catch-up campaigns are underway,^12^ routine immunisation coverage among children remains lower than pre-pandemic levels.^13^

The impact of the pandemic on demand for childhood vaccines (rather than practical barriers) remains poorly understood. While the decline in routine immunisations in the UK and globally is well-documented, there is limited evidence on how the pandemic influenced demand for vaccines other than the Covid-19 vaccine. Several factors suggest that the pandemic may have affected broader vaccine hesitancy. Increased public awareness of immunity, disease transmission, and vaccine development might have boosted parental confidence in vaccines. However, vaccine fatigue,^14^ misinformation,^15^ and potential backfire effects — such as psychological reactance^16^ to vaccine certification policies in the UK^17,18^ and elsewhere^19,20^ — may have contributed to Covid-19 vaccine hesitancy, with possible spillover effects on other vaccines.^21^ Precedents for such spillover exist: after the 2017 Dengvaxia controversy in the Philippines, vaccine confidence dropped from 82% in 2015 to just 22% in 2018,^22^ leading to a substantial decline in routine childhood immunisations. MMR vaccine uptake, for example, fell by over 30 percentage points, while measles cases surged by 2,000% compared to 2017. ^23^

A handful of studies have examined changes in attitudes toward childhood vaccines during the Covid-19 pandemic, most of which have found declines in vaccine confidence, though the evidence in the UK is limited. In the US, research indicates decreased vaccine confidence in specific populations,^24,25^ with Shah et al. (2022) reporting that from 2020 to 2022, more parents believed childhood vaccines could cause illness, death, or harmful side effects in a longitudinal, nationally representative study.^26^ Similarly, declines in vaccine confidence have been observed in Italy^27^ and Turkey.^28^ However, Higgins et al. found no overall change in parental vaccine hesitancy during the pandemic in the US,^29^ while data from Finland suggested a positive spillover effect, with more adults viewing vaccines favourably.^30^ At a global scale, UNICEF’s *State of the World’s Children 2023* report highlights a drop in vaccine safety perceptions, especially among younger people.^31^ However, as with other surveys using a vaccine-agnostic confidence scale, it remains unclear whether this decline reflects a genuine drop in confidence across all vaccines or if it is driven primarily by attitudes toward the Covid-19 vaccine. In a global study involving over 23,000 respondents from 23 countries, Lazarus et al. found that 23.1% of respondents were less willing to get vaccinated for diseases other than Covid-19, while 60.8% were more willing.^32^ In the UK, a UKHSA survey of 1,000 parents found that 27% felt “more positive about routine childhood vaccines” after the Covid-19 vaccination campaign, while 12% felt less positive.^33^ However, such findings should be interpreted with caution, as asking about changes in attitude may lead to “response substitution,” where participants report their current attitude rather than an actual change.^34^ A 2024 pre-print by de Figueiredo et al. also noted a decline in the intent to receive seasonal influenza vaccines among the UK adult population, particularly in some Asian communities.^35^

The impact of the pandemic on attitudes toward childhood vaccines in the UK remains unclear. This study aims to quantify changes in parental intent regarding childhood vaccinations in the UK during the pandemic using three cross-sectional surveys conducted in 2020, 2021, and 2022. Rather than relying on confidence scales—which may be influenced by the introduction of the Covid-19 vaccine—or asking about changes in attitude, this study focuses on cross-sectional measures of vaccine refusal. This study also examines the demographic and socioeconomic factors influencing vaccination intent and how these have evolved since before the pandemic. Understanding these shifts will provide insights into the pandemic’s impact on national and sub-national demand for childhood vaccines.

## Methods

### Data

This study uses data from three cross-sectional online surveys of UK adults. The first was conducted in September and October 2020, before the Covid-19 vaccine rollout, followed by surveys in October 2021 and July-August 2022. In each survey, panel quotas were set based on UK national distributions of age, sex, and region to ensure a nationally representative sample. The 2020 and 2021 surveys examined vaccination beliefs among UK adults (aged 18 and over), each with about 17,000 respondents. The 2022 survey focused exclusively on individuals responsible for making vaccination decisions for children. In the first two surveys, this responsibility was identified through the question: “Are you responsible for decisions relating to the vaccination of children?” Respondents who answered affirmatively were included in the final sample, along with all respondents from the 2022 survey. The total sample size across all surveys is 14,280, with 5,288 respondents in 2020, 4,994 in 2021, and 3,988 in 2022.

Three response variables (RVs) are used to measure parental or caregiver attitudes toward childhood immunisations. All respondents are asked, “Are there any vaccines you have not, or would not, allow your child(ren) to have?” (RV1), with three response options: “yes,” “no,” and “do not know.” Respondents who did not answer “no” were then asked, “Which vaccines, if any, do you have concerns about?” (RV2), where they could select one or more of the following options: “measles, mumps, and rubella (MMR) combined vaccine,” “influenza vaccine,” “human papillomavirus (HPV) vaccine,” and “other (please specify).” In the 2021 and 2022 surveys, “Covid-19 vaccine” was added to this list, reflecting the rollout of Covid-19 vaccines for children.

In the 2022 survey, an additional question was introduced to explore changes in parental or caregiver attitudes toward vaccination: “Since the beginning of the Covid-19 pandemic, has your attitude towards giving your child(ren) routine vaccines changed? (The Covid-19 vaccine is not considered a routine vaccine).” Respondents answered on a three-point scale: “yes,” “no,” or “do not know.” Those who selected “yes” were then asked, “How have your attitudes towards other vaccines for your child(ren) changed?” and provided with a four-point scale ranging from “I feel much more positive about giving my child vaccines” to “I feel much less positive about giving my child vaccines.” These two questions were combined to create RV3, which consolidates responses into three categories: “more positive,” “no change,” and “less positive.” This two-step questioning approach may help mitigate issues with asking directly about changes in sentiment over time.^36^

Individuals’ sex, age, ethnicity, religious affiliation, employment status, income, work status, first language, and region of residence in the UK are also collected and used as explanatory variables to identify the socioeconomic predictors of the response variables. The first level of the Nomenclature of Territorial Unit for Statistics (NUTS1)^37^ is used for regional classification. The respondent’s own Covid-19 vaccine status was also included in the third survey and is used to explore how attitudes vary according to Covid-19 vaccination status. All study variables (response and predictors) are provided in table 1. Supplementary materials table 1 shows the number and percentage of each socio-demographic response variable recorded across all three surveys.

**Table 1.** Study response and predictor variables. All response (RV1-3) and predictor variables are outlined along with variable value options and recodes, as well as the baseline group used for the logistic regression models (see Methods).

| Variable type | Survey Question | Values | Baseline |
| --- | --- | --- | --- |
| **RV1** | Are there any immunisations you have not, or would not, allow children to have? | yes, no, do not know (no) | n/a (response variable) |
| **RV2** | Which vaccines, if any, do you have concerns about? (Select all that apply) | MMR, HPV, influenza, Covid-19 (2021 and 2022 only), other (please specify), none of the above. | n/a (response variable) |
| **RV3**<br>(2022 only) | Since the beginning of the COVID-19 pandemic, have your attitudes towards giving your child(ren) routine vaccines changed? | Responses to "Since the beginning of the Covid-19 pandemic, has your attitude towards giving your child(ren) routine vaccines changed? (The Covid-19 vaccine is not considered a routine vaccine)" and "How have your attitudes towards other vaccines for your child(ren) changed?" recoded to "more positive," "no change," and "less positive." (see SM Questionnaire 2022). | n/a (response variable) |
| **Sex** | I am... | male, female, other (removed from analysis due to low sample size) | female |
| **Religion** | Do you consider yourself... | Atheist/agnostic, Christian, Buddhist (recoded to "other"), Hindu, Muslim, Jewish (recoded to "other"), other, do not wish to answer. | Christian |
| **Education** | What is the highest level of education you have completed? (Select the response that best applies) | 0-4 GCSEs, O-levels, or equivalent (recoded to "level 1-3"); 2+ A levels or equivalents (recoded to "level 1-3"); 5+ GCSEs, O-levels, 1 A level, or equivalents (recoded to "level 1-3"); Undergraduate postgraduate degree or other professional qualification (recoded to "level 4"); apprenticeship (recoded to "none/other"); do not know (recoded to "none/other"); do not wish to answer; other (e.g., vocational, foreign qualifications) (recoded to "none/other") | level 1-3 |
| **Age** | How old are you? | Integer categorised into 18-24, 25-34, 35-44, 45-54, or 55+ | 35-44 |
| **Region** | Which UK region do you live in? | East Midlands, East of England, Greater London, North West, North West, Northern Ireland, Scotland, South East, South West, Wales, West Midlands, Yorkshire and the Humber, Other (removed from data) | South East |
| **Ethnicity** | Which best describes your ethnicity? (Select the response that best applies) | Asian or Asian British: Chinese (recoded to "Asian/Asian British"); Asian or Asian British: Indian (recoded to "Asian/Asian British"); Asian or Asian British: Other (recoded to "Asian/Asian British"); Asian or Asian British: Pakistani (recoded to "Asian/Asian British"); Black, African, Caribbean, or Black British (recoded to "Black/Black British"); White and Asian or White and Asian British (recoded to "Mixed"); White and Black African (recoded to "Mixed"); White and Black Caribbean (recoded to "Mixed"); White: English/Welsh/Scottish/Northern Irish/British (recoded to "White"); White: Irish (recoded to "White"); White: other white background (recoded to "White"); Gypsy or Irish traveller (recoded to "other"); Roma (recoded to "other"); Other; Do not wish to answer. | White |
| **Income** | What is your total household income in GBP (£) from all sources before tax? | Under £15,000; £15,000 - £24,999; £25,000 - £34,999; £35,000 - £44,999; £45,000 - £54,999; £55,000 - £64,000; £65,000 - £99,999; Over £100,000; do not wish to answer | £25,000 - £34,999 |
| **Work status** | Which of the following best describes your work status 6 months ago? | working full-time (including self-employed); working part-time (including self-employed); unemployed; student; looking after the home; unable to work (e.g. short or long-term disability) (recoded to "retired/disabled"); retired (recoded to "retired/disabled"); Do not wish to answer. | working full-time |
| **Language** | What is your first language? | English or Welsh; Polish (recoded to "other"); Punjabi (recoded to "other"); Punjabi (recoded to "other"); Urdu (recoded to "other"); Bengali (recoded to "other"); other; Do not wish to answer. | English or Welsh |
| **Covid-19 vaccine status**<br>(2022 only) | Which of the following best describes how many COVID-19 vaccines you have currently received? | None, one, two, at least 3 doses | two plus a booster |

### Time-varying trends in vaccine refusal

Binomial logistic regressions are used to obtain the predicted probability (converted to population percentages) of vaccine refusal (RV1) for each respondent in the combined dataset. To generate uncertainty around national and regional refusal rate estimates, a total of 100 bootstrap samples of these probabilities are generated. National and regional level refusal rate estimates (and corresponding confidence intervals) are then obtained by multiplying bootstrapped individual refusal probabilities by inverse proportional weights (IPWs) and summing over all individuals in each survey year. In the absence of population-level census data for adults responsible for vaccinating children — data that would allow for post-stratification — we use IPWs to balance socio-demographic characteristics across the three survey waves. This IPW approach assumes that there are no unobserved confounders. This method helps ensure consistent demographic representation across samples, minimising selection bias and enabling valid comparisons over time, assuming no significant changes in the UK demographic structure between 2020 and 2022. In addition to these bootstrapped time-varying estimates of refusal, odds ratios of association (with baselines specified in table 1) between RV1 and predictor variables are obtained. The same approach as described above is used to generate time-varying estimates and confidence intervals for each of the individual vaccines in RV2 and to generate national estimates and socio-demographic determinants. No evidence of strong multicollinearity was found for the socio-demographic predictors in any model as per the adjusted generalised standard error inflation factor (aGSIF),^38^ with all aGSIFs smaller than 1.6. To account for that the question used to form RV1 (“Are there any immunisations you have not, or would not, allow children to have?”) involves the recode of “Don’t know” to “No” in the main analysis, a sensitivity analysis where these uncertain responses are excluded from the analysis is also conducted.

### Self-reported changes in attitudes to childhood vaccines

A proportional odds ordinal logistic regression ^39^ is used to determine the socio-demographic predictors of self-reported change in attitudes towards childhood routine immunisations over the pandemic (RV3). While all socio-demographic predictors in table 1 are included in the model, individual self-reported Covid-19 vaccine status is also included to further control for possible confounding on baseline attitudes when asking about change.^34^

### Model selection

Stepwise model selection using the Akaike Information Criterion (AIC) was performed to identify an appropriate form of each logistic regression model used in the analysis outlined above with interaction terms between survey year and socio-demographic variable permitted to explore directional changes in vaccine refusal within socio-demographic groups over time. Although stepwise AIC selection does not guarantee the optimal model, it provides a systematic way to balance model complexity and explanatory power, aiming for a model that better represents the data than simpler alternatives. This approach ensures that the final model captures relevant relationships between socio-demographic factors and vaccine refusal while adjusting for shifts in attitudes over time, minimising the risk of overfitting. The models with the lowest AIC are reported in the main text (see SM tables 2-7).

### Patient and public involvement

No patients or members or the public were involved in setting the research question or the outcome measures, nor were they involved in developing plans for design or implementation of the study.

All statistical analyses were performed in R. Data are available upon reasonable request.

## Results

### Changes in national and regional refusal over time

Nationally, the reweighted predicted percentages of parents/carers refusing childhood vaccines were 17.3% (95% CI: 16.4 to 18.5) in 2020, 14.8% (95% CI: 13.7 to 15.7, *p*<0.001 compared to 2020) in 2021, and 21.1% (95% CI: 19.8 to 22.5, *p*<0.001 compared to 2020) in 2022. A similar trend was observed across most regions (see Figure 1), with a decline in reported refusal from 2020 to 2021, followed by an increase in 2022. Across all three surveys, the likelihood of vaccine refusal was consistently highest in Greater London. Sensitivity analyses yielded the same temporal trends (see SM table 8); though excluding “don’t know” responses generally increased the percentage of respondents reporting vaccine refusal by 2 to 5 percentage points compared to when “don’t know” responses were recoded as “no.” While refusal fell slightly in all regions of the UK in 2021 compared to 2022 before increasing to values in excess of those in 2022, concerns about specific vaccines reveals a contrasting picture (figure 2 and SM table 9). Nationally, the proportion of parents/carers expressing concerns about the MMR and influenza vaccines declined from 2020 to 2021 (MMR: 9.6 to 6.3%, *p*<0.001; influenza: 12.6 to 8.2%, *p*<0.001) and again from 2021 to 2022 (MMR: 6.3 to 4.6%, *p*<0.001; influenza: 8.2 to 6.1%, *p*<0.001). This decreasing trend at the national level for these two vaccines was also observed consistently across all regions. For the HPV vaccine, a significant national decrease in reported concerns occurred from 2020 to 2021 (7.1 to 4.8%, *p*<0.001), followed by a smaller, non-significant decline from 2021 to 2022 (4.8 to 4.5%, *p*>0.05). Across all three of these vaccines, levels of concern were again highest in Greater London across all survey years (figure 2). For the Covid-19 vaccine, however, the number of parents who reported concerns rose significantly between 2021 and 2022 at the national level (12.4 to 23.0%, *p*<0.001) as well as for all regions.

**Figure 1.**
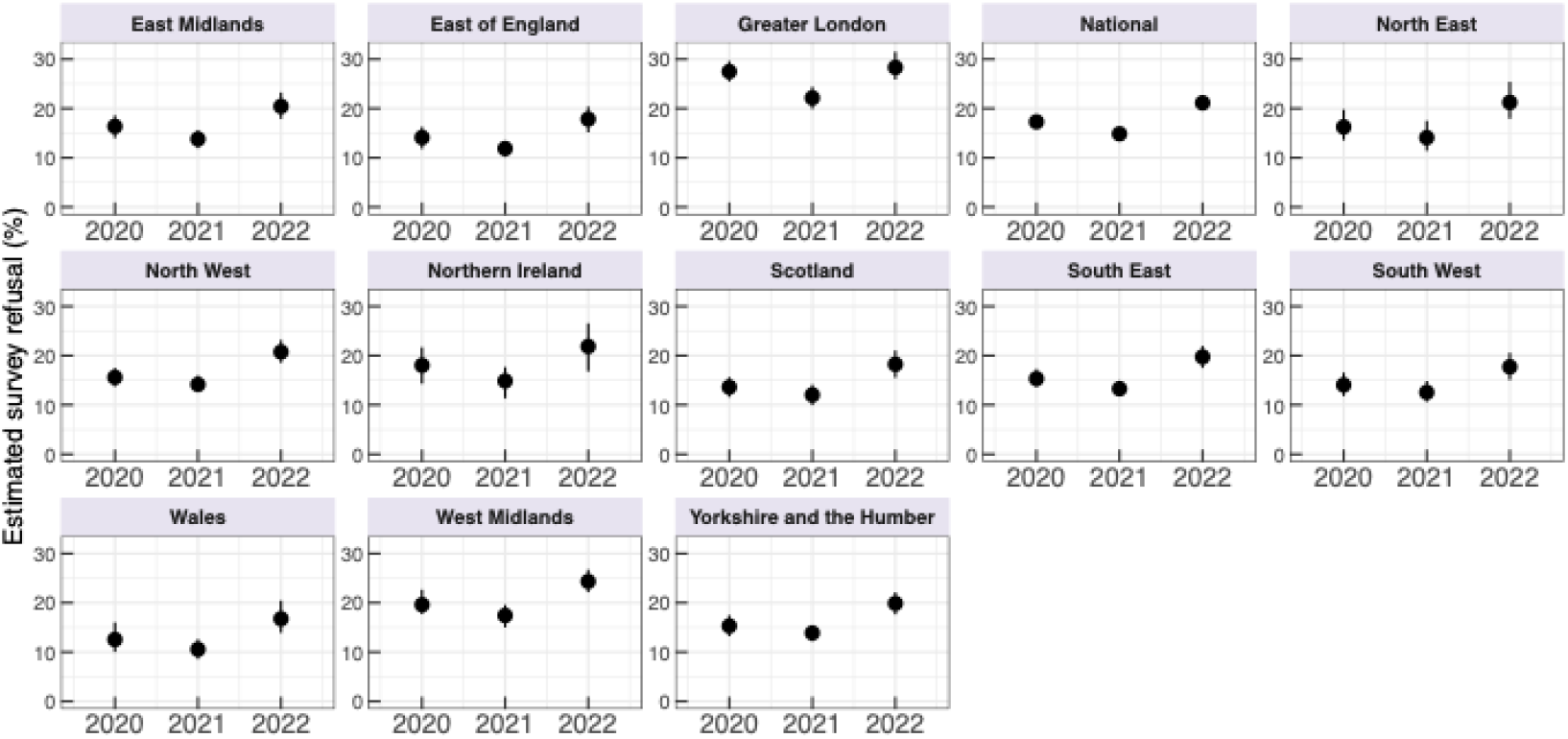
Temporal trends in vaccine refusal across the UK reveal increasing rates of refusal to any vaccine administered to children between 2020 and 2022. Estimated percentage of parents in the UK at the national level and each NUTS1 level with 95% bootstrapped confidence intervals who indicate that there is at least one immunisation they have not, or would not, allow their children to have (RV1).

**Figure 2.**
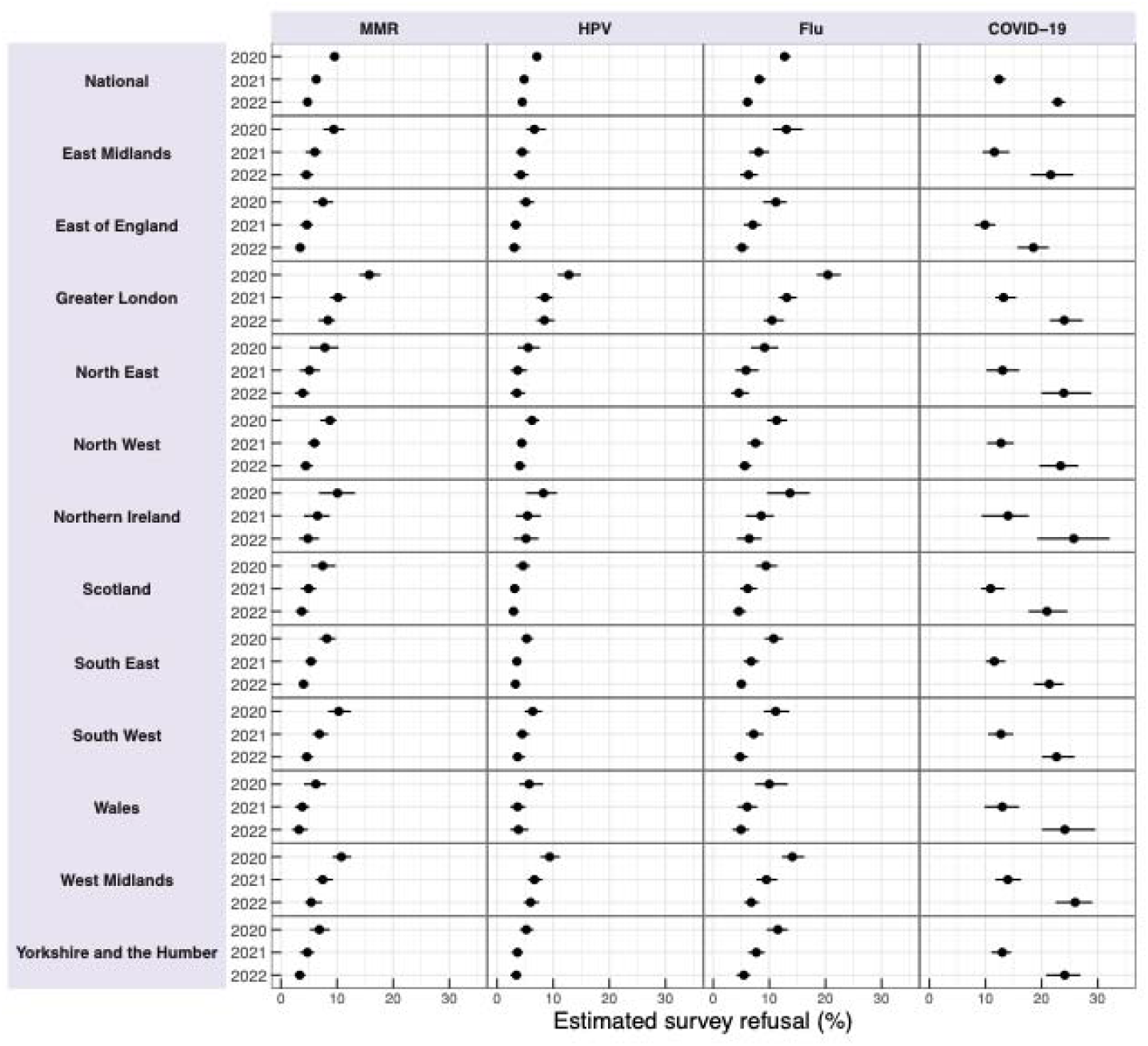
National and regional concern with MMR, HPV, and influenza vaccines has decreased between 2020 and 2022, with concerns increasing markedly for the Covid-19 vaccines over the same period. Estimated percentage of parents in the UK at the national level with 95% bootstrapped confidence intervals and each NUTS1 level who indicate concerns about a specific vaccine (RV2).

### Socio-demographic determinants of vaccine refusal over time

The results of the logistic regression exploring the social, economic and demographic determinants of vaccine refusal (RV1) are shown in figure 3 (odds ratio values are provided in SM table 10 and odds ratio for the sensitivity recoding are provided in SM table 11). An OR larger than one indicates an association with higher refusal. Several socio-demographic factors are associated with vaccine refusal.

**Figure 3.**
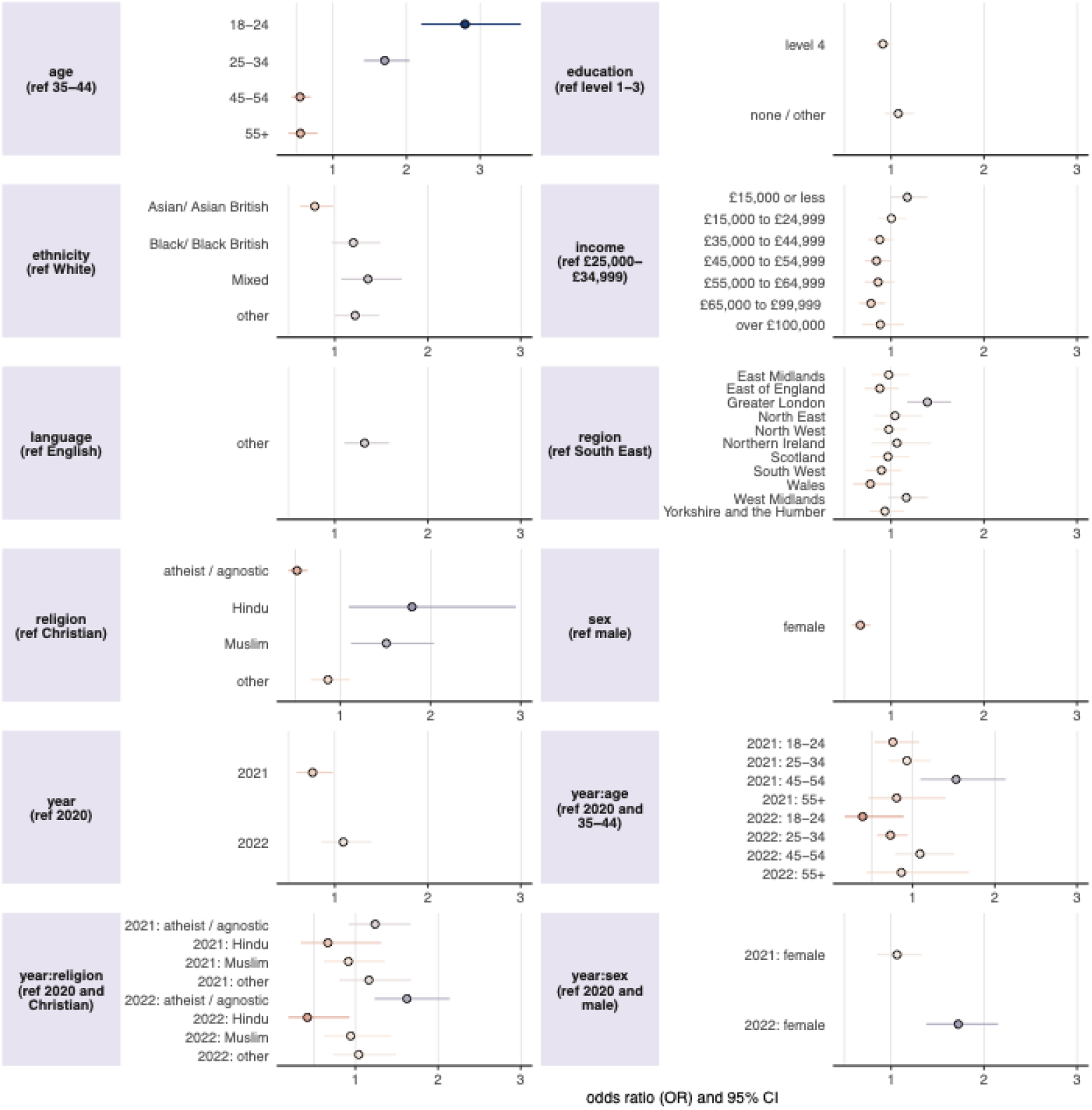
Socio-demographic determinants of vaccine refusal reveal an increase in vaccine refusal among females in 2022 compared to 2020 as well as higher odds of refusal among those in Greater London as well as Hindu and Muslim respondents. The results of the logistic regression of the impact of socio-demographic on the odds of respondents reporting that there is a vaccine they have or would refuse for their child(ren) (RV1). Odds ratios larger than one indicate higher odds of refusal compared to the reference category. Interaction effects are denoted with a colon “:”. The response category “do not wish to answer” – while included in the regression – have been removed from the figure to improve figure visual clarity but can be found in SM table 10.

Female respondents were considerably less likely to report refusal than male respondents throughout the three survey years (odds ratio 0.67, 95% confidence interval 0.58 to 0.78), though this gap appears to have narrowed since 2020, with a positive interaction term between age and sex for 2022 (1.72, 1.38 to 2.15). Compared to the reference age group (35–44 years), younger age groups have higher odds of reporting vaccine refusal: the 18–24 age group has an odds ratio (OR) of 2.79 (95% CI: 2.20 to 3.55), while the 25–34 age group has an odds ratio of 1.70 (95% CI: 1.42 to 2.04). In 2022, compared to 2020, 18–24 and 25–34-year-olds have become less likely to report refusal (0.39, 0.17 to 0.89 and 0.73, 0.57 to 0.94, respectively). Older age groups are found to have much lower odds of refusal, compared to those aged 35–44 years (45– 54: OR 0.55, 0.44 to 0.70; 55 and over: OR 0.56, 0.39 to 0.79). Respondents with Asian or Asian British ethnicities were found to have lower rates of refusal than White respondents (0.78, 0.63 to 0.98), while those reporting Black or Black British or Mixed ethnicities having marginally higher rates of refusal than White respondents (1.20, 0.97 to 1.49 and 1.36, 1.07 to 1.72, respectively). However, Hindu and Muslim respondents were more likely to report refusal compared to Christians (1.78, 1.10 to 2.90 and 1.51, 1.12 to 2.04, respectively), with atheists / agnostics having lower rates of refusal than Christian respondents (0.52, 0.43 to 0.64). However, in 2022, atheists / agnostics report are more likely to report refusal than in 2020 compared to Christians (1.62, 1.23 to 2.1), while Hindus are less likely (0.42, 0.19 to 0.93), with no change detected for Muslim respondents.

Those reporting a first language other than English were more likely to report refusal than those with English as a first language (1.32, 1.10 to 1.58). Higher levels of education (level-4) is found to be slightly more predictive of lower rates of reported refusal than those with level 1-3 education levels (0.91, 0.82 to 1.01) and higher incomes were also associated with lower rates of refusal. Consistent with findings from regional trends in refusal in figure 1, refusal rates in 2021 are lower than 2020 while individuals living in Greater London reported higher rates of refusal than those in the baseline category (South East), with individuals in the West Midlands also reporting slightly higher refusal rates.

### Self-reported changes in attitude to childhood immunisations

Overall, 84.3% of parents reported that their attitudes towards giving their children routine childhood vaccines had not changed (or they were unsure) since the beginning of the pandemic, while 9.2% reported that they had become more positive, while 6.5% had become less positive (unweighted values reported). The results of the ordinal logistic regression evaluating the determinants of the direction of change to changes in perceptions to routine childhood vaccines since the beginning of the pandemic (RV3) are shown in figure 4 and SM table 12.

**Figure 4.**
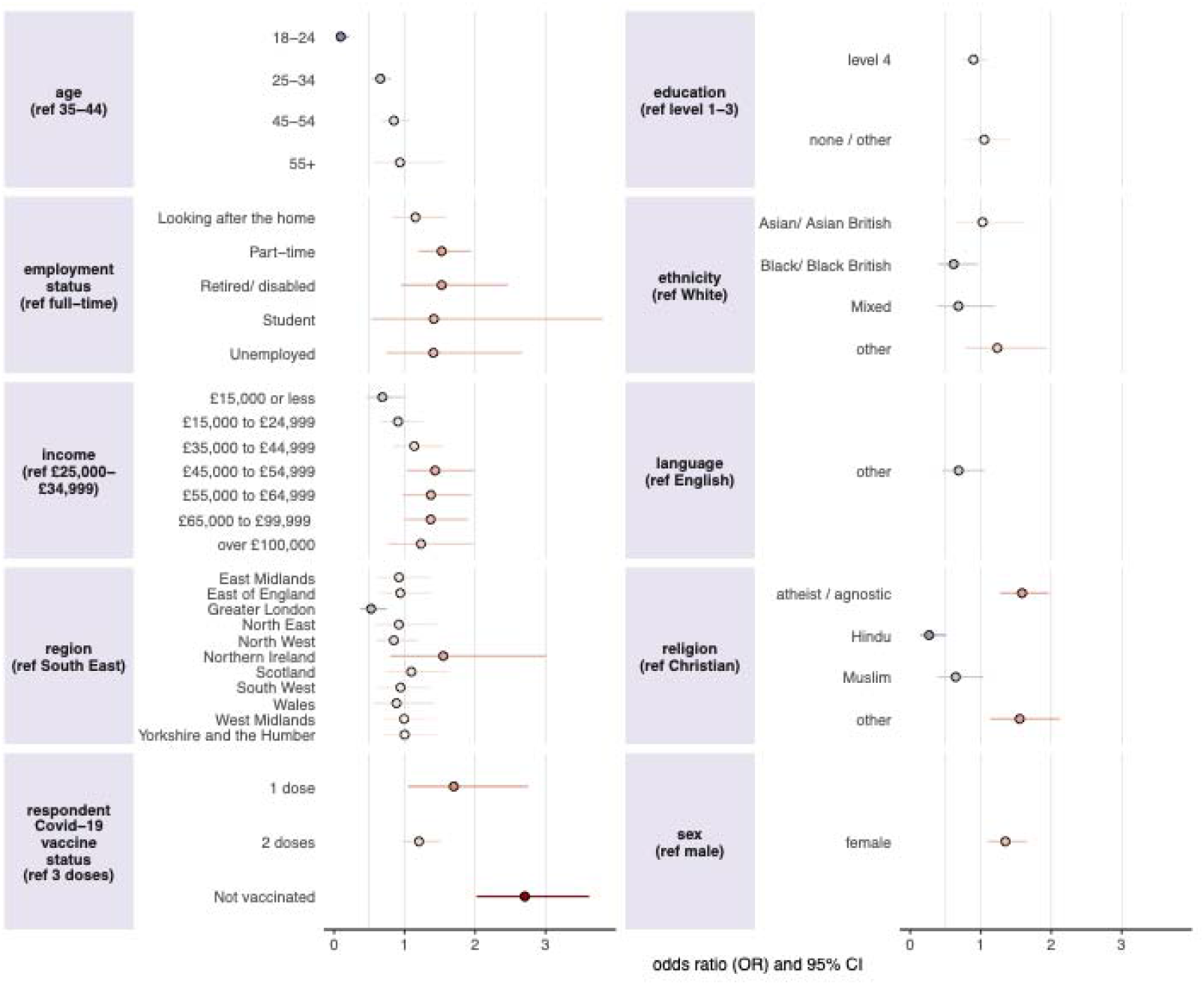
Socio-demographic determinants of changes in parental perceptions towards giving their children routine vaccines since the start of the Covid-19 pandemic. Parents who have not received at least three doses of a Covid-19 vaccine report being less positive in giving their children routine vaccines since the beginning of the Covid-19 pandemic compared to those who have received at least three doses. Younger age groups, those living in Greater London, Black / Black British respondents, and Hindu respondents report a higher likelihood of being more positive about giving their children routine vaccines since the start of the pandemic, relative to their reference categories.

The strongest predictor of an increased negative perception towards routine childhood immunisations was found to be not being vaccinated with a Covid-19 vaccine (2.71, 2.02 to 3.62), with those without at least three doses also exhibiting increased negative perception (one dose: 1.68, 1.05 to 2.75; two doses: 1.20, 0.97 to 1.50). Working part-time (1.53, 1.20 to 1.94), being atheist / agnostic (1.59, 1.28 to 1.97), female (1.35, 1.10 to 1.66), and earning a middling salary were all also associated with an increase in negative perception compared to those working full-time, being Christian, males, and a salary between £25,000 to £34,999, respectively. The strongest predictors of having become more positive about routine childhood vaccines were being aged 18–24 years (0.09, 0.04 to 0.21), being aged 25–34 (0.66, 0.53 to 0.81), living in Greater London (0.53, 0.37 to 0.75), and Hindu respondents (0.27, 0.14 to 0.51). Black / Black British respondents also report more positive attitudes to giving their children vaccines compared to White respondents (0.62, 0.40 to 0.96).

## Discussion

This study provides an in-depth examination of UK parental attitudes toward childhood vaccinations during the Covid-19 pandemic, capturing shifts in refusal rates and identifying key socio-demographic determinants of vaccine refusal and changing attitudes towards childhood vaccines. Our findings reveal a nuanced picture: while overall refusal rates for childhood vaccines initially decreased between 2020 and 2021, they rose again in 2022, surpassing 2020 levels. In contrast, specific concerns about MMR, HPV, and influenza vaccines decreased consistently from 2020 to 2022, suggesting a divergence between general and specific vaccine attitudes, that was explained by increased concerns around Covid-19 vaccines for children. The decreased stated concerns for MMR, HPV, and influenza vaccines are in contrast to observed coverage rates for these vaccines, all of which remain below pre-pandemic levels.^40–42^

Refusal rates were persistently higher in Greater London than in other UK regions. However, London’s 2022 survey responses suggest a relatively positive shift in vaccine attitudes, aligning with findings from the Office for National Statistics on decreasing Covid-19 vaccine hesitancy among Londoners.^43^ This regional trend underscores the importance of context-specific public health messaging and interventions.

Analysis of socio-demographic predictors of refusal (RV1) reveals that respondents who were younger, male, Hindu, Muslim, Black / Black British or spoke a first language other than English were more likely to report vaccine refusal than their respective reference groups across the three years of data. Conversely, higher education levels, higher income and Asian / Asian British respondents were less likely to report refusal. Interestingly, the two youngest age groups, Hindu and Black / British respondents, reported a positive change in attitudes to childhood vaccines over the Covid-19 pandemic. By far the strongest predictor of an increased negative sentiment to childhood vaccines was respondents who had not received at least three doses of Covid-19 vaccines, suggesting a negative spillover effect among individuals who were not fully vaccinated against the Covid-19 vaccine. Overall, however, a small net increase in perceptions towards childhood vaccines was found, a result supported by data from an online survey carried out by the UK Health Security Agency in 2023 of 1,000 parents and 1,000 teenagers, which found no evidence that the Covid-19 vaccination programme had adversely impacted on attitudes to routine vaccines.^33^

### Study limitations

This study provides novel insights into shifts in parental confidence in routine childhood vaccines throughout the pandemic in the UK. However, the definition of vaccine refusal (RV1) is quite broad, capturing both past and future intent, which prohibits making claims about future behaviour. Moreover, the study does not explore reasons behind past or anticipated refusals, so the underlying causes for these trends cannot be inferred from the current data. Additionally, the introduction of additional vaccines (in this case, Covid-19) to a multiple-choice battery (as used in RV2) may induce survey bias into responses for other vaccines, which has the possibility of downwardly biasing estimates for concerns around other vaccines.

Although it’s possible that the discrepancy could stem from another routine childhood vaccine not explicitly prompted in the survey, there was no indication of this from responses provided under the “Other” option in free text. While the question did specify “routine childhood vaccines,” some respondents may have interpreted this as including the Covid-19 vaccine, thus influencing their responses.

## Conclusion

This study provides valuable insights into the evolving dynamics of parental and caregiver confidence in routine childhood vaccines in the UK during the COVID-19 pandemic. While vaccine refusal rates generally increased over the study period, the observed decrease in concerns about MMR, HPV, and influenza vaccines suggests that primary concerns in 2022 were directed at Covid-19 vaccines for children. The marked negative shift in attitudes towards childhood vaccines among respondents who had not received at least three doses of a COVID-19 vaccine highlights a potential spillover effect of Covid-19 vaccine hesitancy onto routine childhood vaccines, and further research could focus on exploring hesitancy concerns in this cohort. The recent declines in MMR vaccine uptake across the UK do not appear to be linked to parental confidence in these vaccines, as indicated by this study’s findings. Further research is essential to understand the current drivers and barriers to childhood immunisations to address the decade-long slide in uptake.

## Data Availability

All data produced in the present study are available upon reasonable request to the authors

## ETHICS APPROVAL

Survey data collection from 2020 was approved by the Imperial College Research Ethics Committee on 24 July 2020 with reference 20IC6133. Survey data from 2021 was approved on 7 April 2021 with reference 25637. Survey data collection from 2022 was approved on 31 March 2022 by the London School of Hygiene & Tropical Medicine’s Ethics Committee with reference 26854.

