## supplementary material for "Changing parental perceptions to childhood immunisations during the Covid-19 pandemic in the UK: evidence from three cross-sectional surveys between 2020 and 2022"

**Supplementary materials for Trends in self-reported parental refusal of childhood immunisations during the Covid-19 pandemic in the UK: evidence from three cross-sectional surveys between 2020 and 2022**

Rosie Thistlethwayte^1^ and Alexandre de Figueiredo^1,2†^

**Table 1: number of responses by socioeconomic/ demographic group within each survey**

|  | 2020 (%)  N = 5288 |  | 2021 (%)  N = 4994 |  | 2022 (%)  N = 3988 |  |
| --- | --- | --- | --- | --- | --- | --- |
|  | **N** | **%** | **N** | **%** | **N** | **%** |
| Sex |  |  |  |  |  |  |
| Female | 2821 | 53.3 | 3074 | 61.6 | 2495 | 62.6 |
| Male | 2467 | 46.7 | 1924 | 38.5 | 1496 | 37.5 |
| Age group |  |  |  |  |  |  |
| 18-24 | 444 | 8.4 | 435 | 8.7 | 34 | 0.9 |
| 25-34 | 1357 | 25.7 | 1309 | 26.2 | 1168 | 29.3 |
| 35-44 | 1829 | 34.6 | 1726 | 34.6 | 1730 | 43.4 |
| 45-54 | 1230 | 23.3 | 1141 | 22.8 | 934 | 23.4 |
| 55+ | 428 | 8.1 | 387 | 7.7 | 125 | 3.1 |
| Region |  |  |  |  |  |  |
| East Midlands | 378 | 7.1 | 332 | 6.6 | 317 | 7.9 |
| East of England | 447 | 8.5 | 415 | 8.3 | 315 | 7.9 |
| London | 799 | 15.1 | 739 | 14.8 | 464 | 11.6 |
| North East | 220 | 4.2 | 209 | 4.2 | 189 | 4.7 |
| North West | 606 | 11.5 | 567 | 11.4 | 479 | 12.0 |
| Northern Ireland | 160 | 3.0 | 152 | 3.0 | 81 | 2.0 |
| Scotland | 404 | 7.6 | 381 | 7.6 | 310 | 7.8 |
| South East | 744 | 14.1 | 647 | 13.0 | 557 | 14.0 |
| South West | 359 | 6.8 | 373 | 7.5 | 309 | 7.7 |
| Wales | 202 | 3.8 | 224 | 4.5 | 181 | 4.5 |
| West Midlands | 506 | 9.6 | 530 | 10.6 | 410 | 10.3 |
| Yorkshire and the Humber | 463 | 8.8 | 425 | 8.5 | 376 | 9.4 |
| Education |  |  |  |  |  |  |
| Level 1-3 | 2144 | 40.5 | 2121 | 42.5 | 1642 | 41.2 |
| Level 4 | 2495 | 47.2 | 2295 | 46.0 | 1913 | 48.0 |
| None/ other | 618 | 11.7 | 536 | 10.7 | 416 | 10.4 |
| Do not wish to answer | 31 | 0.6 | 44 | 0.9 | 19 | 0.5 |
| Work status |  |  |  |  |  |  |
| Full-time | 3060 | 57.9 | 2785 | 55.8 | 2341 | 58.7 |
| Part-time | 1122 | 21.2 | 1130 | 22.6 | 940 | 23.6 |
| Student | 115 | 2.2 | 105 | 2.1 | 33 | 0.8 |
| Unemployed | 195 | 3.7 | 217 | 4.3 | 89 | 2.2 |
| Looking after the home | 545 | 10.3 | 502 | 10.1 | 421 | 10.6 |
| Retired/ disabled | 232 | 4.4 | 237 | 4.7 | 156 | 3.9 |
| Do not wish to answer | 19 | 0.4 | 22 | 0.4 | 11 | 0.3 |
| Religion |  |  |  |  |  |  |
| Atheist/ agnostic | 1378 | 26.1 | 1258 | 25.2 | 1190 | 29.8 |
| Christian | 2543 | 48.1 | 2381 | 47.7 | 1748 | 43.8 |
| Do not wish to answer | 387 | 7.3 | 375 | 7.5 | 341 | 8.6 |
| Hindu | 100 | 1.9 | 118 | 2.4 | 70 | 1.8 |
| Muslim | 302 | 5.7 | 350 | 7.0 | 211 | 5.3 |
| Other | 578 | 10.9 | 516 | 10.3 | 431 | 10.8 |
| Ethnicity |  |  |  |  |  |  |
| Asian/ Asian British | 394 | 7.5 | 469 | 9.4 | 289 | 7.2 |
| Black/ Black British | 181 | 3.4 | 188 | 3.8 | 164 | 4.1 |
| Mixed | 159 | 3.0 | 156 | 3.1 | 99 | 2.5 |
| White | 4122 | 78.0 | 3782 | 75.7 | 3211 | 80.5 |
| Other | 396 | 7.5 | 365 | 7.3 | 209 | 5.2 |
| Do not wish to answer | 36 | 0.7 | 38 | 0.8 | 19 | 0.5 |
| Language |  |  |  |  |  |  |
| English or Welsh | 4746 | 89.8 | 4533 | 90.8 | 3740 | 93.8 |
| Other | 517 | 9.8 | 442 | 8.9 | 239 | 6.0 |
| Do not wish to answer | 25 | 0.5 | 19 | 0.4 | 9 | 0.2 |
| Income |  |  |  |  |  |  |
| Under ¬£15,000 | 525 | 9.9 | 487 | 9.8 | 315 | 7.9 |
| ¬£15,000 to ¬£24,999 | 859 | 16.2 | 777 | 15.6 | 513 | 12.9 |
| ¬£25,000 to ¬£34,999 | 881 | 16.7 | 921 | 18.4 | 666 | 16.7 |
| ¬£35,000 to ¬£44,999 | 807 | 15.3 | 761 | 15.2 | 626 | 15.7 |
| ¬£45,000 to ¬£54,999 | 677 | 12.8 | 638 | 12.8 | 544 | 13.6 |
| ¬£55,000 to ¬£64,999 | 455 | 8.6 | 430 | 8.6 | 429 | 10.8 |
| ¬£65,000 to ¬£99,999 | 613 | 11.6 | 552 | 11.1 | 539 | 13.5 |
| Over ¬£100,000 | 206 | 3.9 | 200 | 4.0 | 170 | 4.3 |
| Do not wish to answer | 265 | 5.0 | 232 | 4.6 | 189 | 4.7 |

**Table 2: Model comparison for RV1: “is there a vaccine you have or would refuse for your child?”**

| **Model** | **Akaike Information Criterion (AIC)** | **Bayesian Information Criterion (BIC)** |
| --- | --- | --- |
| **All variables, no interactions**  Y ~ Year + Age + Sex + Income + Region + WorkStatus + Ethnicity + Education + Religion + Language | 12,701 | 13,064 |
| **Stepwise AIC final model**  Y ~ Age*Year + Sex*Year + Region + Income + Education + Ethnicity + Religion*Year + Language | 12,656 | 13,125 |

**Table 3: Model comparison for RV2: concerns around the MMR vaccine**

| **Model** | **Akaike Information Criterion (AIC)** | **Bayesian Information Criterion (BIC)** |
| --- | --- | --- |
| **All variables, no interactions**  Y ~ Year + Age + Sex + Income + Region + WorkStatus + Ethnicity + Education + Religion + Language | 6,983 | 7,346 |
| **Stepwise AIC final model**  Y ~ Year + Age + Sex + Region + Income + Ethnicity + Religion + Language | 6,977 | 7,272 |

**Table 4: Model comparison for RV2: concerns around the influenza vaccine**

| **Model** | **Akaike Information Criterion (AIC)** | **Bayesian Information Criterion (BIC)** |
| --- | --- | --- |
| **All variables, no interactions**  Y ~ Year + Age + Sex + Income + Region + WorkStatus + Ethnicity + Education + Religion + Language | 8,371 | 8,743 |
| **Stepwise AIC final model**  Y ~ Year + Age + Sex + Region + Education + Ethnicity + Religion + Language | 8,364 | 8,621 |

**Table 5: Model comparison for RV2: concerns around the HPV vaccine**

| **Model** | **Akaike Information Criterion (AIC)** | **Bayesian Information Criterion (BIC)** |
| --- | --- | --- |
| **All variables, no interactions**  Y ~ Year + Age + Sex + Income + Region + WorkStatus + Ethnicity + Education + Religion + Language | 5,941 | 6,304 |
| **Stepwise AIC final model**  Y ~ Year + Age + Sex + Region + Income + Ethnicity + Religion + Language | 5,924 | 6,203 |

**Table 6: Model comparison for RV2: concerns around the Covid-19 vaccine**

| **Model** | **Akaike Information Criterion (AIC)** | **Bayesian Information Criterion (BIC)** |
| --- | --- | --- |
| **All variables, no interactions**  Y ~ Year + Age + Sex + Income + Region + WorkStatus + Ethnicity + Education + Religion + Language | 7,913 | 8,276 |
| **Stepwise AIC final model**  Y ~ Education + WorkStatus + Ethnicity | 7.890 | 8,117 |

**Table 7: Model comparison for RV3 of “Since the beginning of the COVID-19 pandemic, how have your attitudes towards routine childhood vaccines changed?”**

| **Model** | **Akaike Information Criterion (AIC)** | **Bayesian Information Criterion (BIC)** |
| --- | --- | --- |
| **All variables, no interactions**  Y ~ Year + CovidVaccinationStatus + Age + Sex + Income + Region + WorkStatus + Ethnicity + Education + Religion + Language | 4,077 | 4,339 |
| **Stepwise AIC final model**  Y ~ CovidVaccinationStatus + Age + Sex + Income + Region + WorkStatus + Religion + Language | 4,075 | 4,325 |

**Table 8: Predicted percentages of vaccine refusal (“is there a vaccine you have or would refuse for your child?”) for the main analysis (“don’t know” recoded to “no”) and a sensitivity analysis that removes “don’t know” responses.**

| **Region** | **Model predicted percentage and 95% CI (2020)** | | **Model predicted percentage and 95% CI (2021)** | | **2022 Predicted probability**  **(95% CI)** | |
| --- | --- | --- | --- | --- | --- | --- |
|  | **main analysis** | **sensitivity** | **main analysis** | **sensitivity** | **main analysis** | **sensitivity** |
| National | 17.3 (16.4, 18.5) | 19.6 (18.5, 20.6) | 14.8 (13.7, 15.7) | 17.1 (15.9, 18.3) | 21.1 (19.8, 22.5) | 24.1 (22.5, 25.6) |
| East Midlands | 16.4 (14.0, 18.6) | 18.4 (15.0, 21.0) | 13.8 (11.9, 15.6) | 15.7 (13.3, 17.9) | 20.4 (17.9, 23.2) | 23.5 (20.1, 26.5) |
| East of England | 14.1 (11.8, 16.2) | 15.8 (13.1, 18.2) | 11.9 (10.1, 13.3) | 13.8 (11.6, 16.3) | 17.8 (15.1, 20.4) | 20.6 (17.4, 23.7) |
| Greater London | 27.5 (25.5, 29.6) | 31.6 (29.1, 34.1) | 22.1 (19.9, 24.4) | 26.0 (23.7, 29.0) | 28.3 (25.9, 31.5) | 32.0 (28.4, 35.6) |
| North East | 16.2 (13.6, 19.8) | 17.6 (15.0, 20.6) | 14.1 (11.4, 17.5) | 15.5 (12.7, 18.8) | 21.2 (17.9, 25.4) | 24.1 (19.9, 28.5) |
| North West | 15.6 (13.7, 17.5) | 17.9 (16.1, 20.0) | 14.1 (12.6, 16.0) | 16.4 (14.5, 18.8) | 20.7 (18.5, 23.2) | 23.7 (20.9, 26.1) |
| Northern Ireland | 18.0 (14.3, 21.7) | 20.1 (16.3, 24.0) | 14.9 (11.3, 17.7) | 16.6 (13.6, 20.5) | 21.9 (16.8, 26.6) | 24.2 (19.8, 29.2) |
| Scotland | 13.6 (11.6, 15.6) | 15.5 (13.6, 17.9) | 12.0 (9.9, 14.1) | 13.6 (11.6, 15.6) | 18.2 (15.5, 21.0) | 21.1 (18.2, 24.5) |
| South East | 15.3 (13.7, 17.2) | 16.9 (15.0, 19.1) | 13.3 (11.9, 14.9) | 15.1 (13.1, 16.8) | 19.7 (17.5, 22.0) | 22.2 (19.5, 24.7) |
| South West | 14.0 (11.8, 16.5) | 15.8 (13.9, 18.4) | 12.5 (10.6, 14.8) | 14.3 (12.2, 16.6) | 17.7 (15.1, 20.6) | 20.2 (17.3, 22.6) |
| Wales | 12.5 (10.0, 16.0) | 14.0 (10.5, 16.8) | 10.5 (8.6, 12.6) | 12.4 (9.9, 15.4) | 16.8 (13.9, 20.4) | 19.3 (15.0, 23.2) |
| West Midlands | 19.6 (17.7, 22.6) | 23.0 (20.8, 25.8) | 17.4 (15.0, 19.6) | 20.6 (17.9, 22.9) | 24.3 (22.1, 26.7) | 28.3 (25.7, 31.4) |
| Yorkshire and  The Humber | 15.3 (13.2, 17.5) | 17.4 (15.1, 20.0) | 13.9 (12.2, 15.5) | 15.5 (13.4, 17.5) | 19.9 (17.6, 22.1) | 22.7 (20.0, 25.4) |

**Table 9: predicted probability of having concerns about MMR, HPV, Flu or COVID-19 vaccines**

| Region | Year | MMR | HPV | Flu | Covid-19 |
| --- | --- | --- | --- | --- | --- |
| National | 2020 | 9.6 (8.8, 10.4) | 7.1 (6.5, 7.9) | 12.6 (11.7, 13.4) | NA |
| National | 2021 | 6.3 (5.6, 7.0) | 4.8 (4.3, 5.3) | 8.2 (7.6, 9.0) | 12.4 (11.5, 13.3) |
| National | 2022 | 4.6 (3.7, 5.3) | 4.5 (4.0, 5.2) | 6.1 (5.2, 7.0) | 23.0 (21.6, 24.5) |
| East Midlands | 2020 | 9.4 (7.6, 12.0) | 6.5 (5.3, 8.1) | 13 (11.1, 15.2) | NA |
| East Midlands | 2021 | 6.0 (4.8, 7.5) | 4.4 (3.2, 5.8) | 8.2 (6.8, 9.9) | 11.5 (9.3, 13.6) |
| East Midlands | 2022 | 4.4 (3.3, 5.7) | 4.2 (3.2, 5.3) | 6.3 (5.1, 7.8) | 21.5 (17.7, 25) |
| East of England | 2020 | 7.2 (5.8, 9.2) | 5.2 (3.4, 6.8) | 11.1 (9.1, 13.1) | NA |
| East of England | 2021 | 4.5 (3.3, 5.8) | 3.3 (2.2, 4.5) | 7.1 (5.9, 8.6) | 9.9 (8.2, 11.7) |
| East of England | 2022 | 3.2 (2.4, 4.2) | 3.1 (2.1, 4.0) | 5.1 (4.0, 6.3) | 18.6 (15.7, 21.1) |
| Greater London | 2020 | 15.9 (14, 17.7) | 12.9 (11.3, 15.3) | 20.1 (18, 22) | NA |
| Greater London | 2021 | 10.2 (8.9, 11.6) | 8.6 (7.3, 10.3) | 13.1 (11.7, 14.7) | 13.3 (11.5, 15.2) |
| Greater London | 2022 | 8.1 (6.4, 9.4) | 8.6 (7.1, 10.4) | 10.3 (8.7, 12.1) | 24.3 (21.1, 27.5) |
| North East | 2020 | 7.7 (5.5, 10.3) | 5.3 (3.1, 7.8) | 9.2 (6.7, 11.6) | NA |
| North East | 2021 | 5.0 (3.5, 6.7) | 3.5 (2.2, 5.0) | 5.9 (4.4, 7.9) | 13.1 (10.0, 16.6) |
| North East | 2022 | 3.7 (2.5, 4.9) | 3.3 (1.8, 4.8) | 4.7 (3.2, 6.6) | 24.1 (19.0, 29.1) |
| North West | 2020 | 8.8 (7.3, 10.6) | 6.1 (4.8, 7.7) | 11.2 (9.8, 12.6) | NA |
| North West | 2021 | 6.0 (4.9, 7.2) | 4.3 (3.4, 5.3) | 7.5 (6.5, 8.7) | 12.7 (10.8, 14.6) |
| North West | 2022 | 4.4 (3.1, 5.3) | 4.0 (3.1, 5.2) | 5.6 (4.5, 6.9) | 23.4 (20.6, 27.1) |
| Northern  Ireland | 2020 | 10.4 (7, 13.2) | 8.0 (5.5, 11.6) | 13.8 (10.2, 17.3) | NA |
| Northern  Ireland | 2021 | 6.6 (4.3, 8.5) | 5.3 (3.4, 7.6) | 8.8 (6.5, 11.3) | 13.5 (9.8, 16.6) |
| Northern  Ireland | 2022 | 4.8 (3.0, 6.5) | 5.0 (3.1, 7.2) | 6.6 (4.4, 8.6) | 24.8 (18.5, 31.2) |
| Scotland | 2020 | 7.3 (5.6, 9.1) | 4.7 (3.5, 6.3) | 9.5 (7.5, 11.7) | NA |
| Scotland | 2021 | 4.8 (3.6, 6.3) | 3.2 (2.4, 4.4) | 6.3 (5.0, 7.7) | 10.8 (9.0, 13.0) |
| Scotland | 2022 | 3.6 (2.4, 4.9) | 3.0 (2.1, 3.9) | 4.7 (3.6, 5.9) | 20.9 (17.5, 23.9) |
| South East | 2020 | 8.2 (7.0, 9.4) | 5.4 (4.3, 6.7) | 10.7 (9.2, 12.5) | NA |
| South East | 2021 | 5.3 (4.2, 6.4) | 3.6 (2.8, 4.3) | 6.9 (5.8, 8.0) | 11.4 (9.7, 13.1) |
| South East | 2022 | 3.9 (3.0, 4.6) | 3.4 (2.7, 4.3) | 5.0 (4.0, 6.1) | 21.3 (19.1, 24.2) |
| South West | 2020 | 10.3 (7.8, 12.8) | 6.4 (5.1, 8.4) | 10.8 (8.1, 13.1) | NA |
| South West | 2021 | 6.9 (5.4, 8.6) | 4.5 (3.4, 5.7) | 7.0 (5.4, 8.9) | 12.9 (10.6, 15.6) |
| South West | 2022 | 4.5 (3.2, 5.9) | 3.7 (2.9, 5.0) | 4.6 (3.5, 6.2) | 23.2 (19.7, 27.3) |
| Wales | 2020 | 6.3 (4.4, 8.3) | 5.8 (3.7, 8.6) | 10.2 (7.3, 12.7) | NA |
| Wales | 2021 | 3.9 (2.6, 5.4) | 3.7 (2.5, 5.1) | 6.2 (4.3, 7.9) | 13.4 (10.7, 16.8) |
| Wales | 2022 | 3.2 (2.0, 4.5) | 3.9 (2.5, 5.8) | 5.1 (3.3, 6.8) | 24.8 (21.0, 30.2) |
| West Midlands | 2020 | 10.8 (8.9, 12.5) | 9.3 (7.6, 10.9) | 13.9 (11.7, 15.8) | NA |
| West Midlands | 2021 | 7.5 (6.2, 8.8) | 6.6 (5.2, 7.8) | 9.4 (8, 11.0) | 14.0 (12.6, 16.0) |
| West Midlands | 2022 | 5.3 (4.0, 6.6) | 6.0 (4.7, 7.5) | 6.7 (5.5, 8.2) | 26.2 (23.1, 29.9) |
| Yorkshire and  the Humber | 2020 | 6.9 (5.4, 8.5) | 5.3 (4.1, 6.8) | 11.4 (9.1, 13.1) | NA |
| Yorkshire and  the Humber | 2021 | 4.8 (3.8, 6.0) | 3.7 (2.7, 4.8) | 7.7 (6.1, 9.2) | 13.1 (10.8, 15.5) |
| Yorkshire and  the Humber | 2022 | 3.3 (2.4, 4.3) | 3.5 (2.6, 4.5) | 5.4 (4.3, 6.5) | 24.4 (20.5, 28.0) |

**Table 10: Odds of vaccine refusal by socioeconomic/ demographic group (with ‘Don’t Know’ responses recoded to ‘No’**

| **Variable Group** | **Variable** | **OR (95% CI)** | ***p*-value** |
| --- | --- | --- | --- |
| Age group (Ref = 35-44) | 18-24yrs | 2.796 (2.199, 3.553) | 0.000 |
| Age group (Ref = 35-44) | 25-34yrs | 1.703 (1.421, 2.041) | 0.000 |
| Age group (Ref = 35-44) | 45-54yrs | 0.551 (0.436, 0.697) | 0.000 |
| Age group (Ref = 35-44) | 55+yrs | 0.556 (0.392, 0.788) | 0.001 |
| Year (Ref = 2020) | 2021 | 0.760 (0.589, 0.981) | 0.035 |
| Year (Ref = 2020) | 2022 | 1.091 (0.855, 1.393) | 0.483 |
| Sex (Ref = Male) | Female | 0.670 (0.576, 0.778) | 0.000 |
| Region (Ref = South East) | East Midlands | 0.975 (0.792, 1.200) | 0.808 |
| Region (Ref = South East) | East of England | 0.879 (0.714, 1.081) | 0.221 |
| Region (Ref = South East) | Greater London | 1.390 (1.174, 1.645) | 0.000 |
| Region (Ref = South East) | North East | 1.042 (0.814, 1.335) | 0.743 |
| Region (Ref = South East) | North West | 0.975 (0.813, 1.170) | 0.787 |
| Region (Ref = South East) | Northern Ireland | 1.063 (0.791, 1.428) | 0.686 |
| Region (Ref = South East) | Scotland | 0.966 (0.781, 1.195) | 0.751 |
| Region (Ref = South East) | South West | 0.897 (0.724, 1.112) | 0.320 |
| Region (Ref = South East) | Wales | 0.776 (0.592, 1.018) | 0.067 |
| Region (Ref = South East) | West Midlands | 1.164 (0.970, 1.396) | 0.103 |
| Region (Ref = South East) | Yorkshire and the Humber | 0.933 (0.766, 1.137) | 0.492 |
| Income (Ref = £25,000-£34,999) | £15,000 to £24,999 | 1.003 (0.861, 1.169) | 0.965 |
| Income (Ref = £25,000-£34,999) | 35,000 to £44,000 | 0.878 (0.752, 1.026) | 0.101 |
| Income (Ref = £25,000-£34,999) | £45,000 to £54,999 | 0.843 (0.715, 0.993) | 0.041 |
| Income (Ref = £25,000-£34,999) | £55,000 to £64,999 | 0.860 (0.716, 1.034) | 0.108 |
| Income (Ref = £25,000-£34,999) | £65,000 to £99,999 | 0.783 (0.657, 0.934) | 0.007 |
| Income (Ref = £25,000-£34,999) | Do not wish to answer | 0.784 (0.613, 1.003) | 0.053 |
| Income (Ref = £25,000-£34,999) | Over £100,000 | 0.884 (0.689, 1.134) | 0.332 |
| Income (Ref = £25,000-£34,999) | Under £15,000 | 1.174 (0.989, 1.395) | 0.067 |
| Education (Ref = Level 1-3) | Do not wish to answer | 0.751 (0.434, 1.300) | 0.307 |
| Education (Ref = Level 1-3) | Level 4 | 0.912 (0.823, 1.010) | 0.078 |
| Education (Ref = Level 1-3) | None/ other | 1.077 (0.930, 1.246) | 0.324 |
| Ethnicity (Ref = White) | Asian/ Asian British | 0.785 (0.626, 0.984) | 0.036 |
| Ethnicity (Ref = White) | Black/ Black British | 1.199 (0.965, 1.489) | 0.101 |
| Ethnicity (Ref = White) | Do not wish to answer | 0.960 (0.540, 1.708) | 0.891 |
| Ethnicity (Ref = White) | Mixed | 1.356 (1.069, 1.720) | 0.012 |
| Ethnicity (Ref = White) | Other | 1.217 (1.003, 1.478) | 0.047 |
| Religion (Ref = Christian) | Atheist/ agnostic | 0.520 (0.425, 0.637) | 0.000 |
| Religion (Ref = Christian) | Do not wish to answer | 1.018 (0.772, 1.343) | 0.898 |
| Religion (Ref = Christian) | Hindu | 1.795 (1.095, 2.944) | 0.020 |
| Religion (Ref = Christian) | Muslim | 1.510 (1.119, 2.039) | 0.007 |
| Religion (Ref = Christian) | Other | 0.863 (0.675, 1.105) | 0.242 |
| First Language (Ref = English) | Do not wish to answer | 1.941 (1.029, 3.662) | 0.040 |
| First Language (Ref = English) | Other | 1.320 (1.101, 1.582) | 0.003 |
| Interaction: Year*Age | 2021: 18-24yrs | 0.760 (0.535, 1.081) | 0.127 |
| Interaction: Year*Age | 2021: 25-34yrs | 0.931 (0.711, 1.219) | 0.604 |
| Interaction: Year*Age | 2021: 45-54yrs | 1.529 (1.095, 2.133) | 0.013 |
| Interaction: Year*Age | 2021: 55+yrs | 0.806 (0.463, 1.401) | 0.443 |
| Interaction: Year*Age | 2022: 18-24yrs | 0.392 (0.173, 0.890) | 0.025 |
| Interaction: Year*Age | 2022: 25-34yrs | 0.730 (0.568, 0.940) | 0.015 |
| Interaction: Year*Age | 2022: 45-54yrs | 1.090 (0.790, 1.504) | 0.600 |
| Interaction: Year*Age | 2022: 55+yrs | 0.864 (0.443, 1.686) | 0.668 |
| Interaction: Year*Sex | 2021: Female | 1.064 (0.852, 1.329) | 0.584 |
| Interaction: Year*Sex | 2022: Female | 1.723 (1.379, 2.152) | 0.000 |
| Interaction: Year*Religion | 2021: Atheist/ agnostic | 1.239 (0.921, 1.667) | 0.156 |
| Interaction: Year*Religion | 2022: Atheist/ agnostic | 1.624 (1.233, 2.140) | 0.001 |
| Interaction: Year*Religion | 2021: Do not wish to answer | 1.097 (0.735, 1.638) | 0.650 |
| Interaction: Year*Religion | 2022: Do not wish to answer | 0.783 (0.523, 1.174) | 0.237 |
| Interaction: Year*Religion | 2021: Hindu | 0.668 (0.340, 1.310) | 0.240 |
| Interaction: Year*Religion | 2022: Hindu | 0.420 (0.190, 0.928) | 0.032 |
| Interaction: Year*Religion | 2021: Muslim | 0.915 (0.619, 1.353) | 0.658 |
| Interaction: Year*Religion | 2022: Muslim | 0.944 (0.619, 1.440) | 0.788 |
| Interaction: Year*Religion | 2021: Other | 1.164 (0.810, 1.673) | 0.412 |
| Interaction: Year*Religion | 2022: Other | 1.040 (0.726, 1.489) | 0.830 |

**Table 11: Odds of vaccine refusal by socioeconomic/ demographic group (sensitivity analysis: ‘don’t know’ responses excluded)**

| **Variable Group** | **Variable** | **OR (95% CI)** | ***p*-value** |
| --- | --- | --- | --- |
| Age group (Ref = 35-44) | 18-24yrs | 2.856 (2.221, 3.674) | 0.000 |
| Age group (Ref = 35-44) | 25-34yrs | 1.666 (1.384, 2.005) | 0.000 |
| Age group (Ref = 35-44) | 45-54yrs | 0.553 (0.436, 0.702) | 0.000 |
| Age group (Ref = 35-44) | 55+yrs | 0.542 (0.381, 0.771) | 0.001 |
| Year (Ref = 2020) | 2021 | 0.760 (0.586, 0.987) | 0.039 |
| Year (Ref = 2020) | 2022 | 1.065 (0.830, 1.366) | 0.623 |
| Sex (Ref = Male) | Female | 0.619 (0.531, 0.722) | 0.000 |
| Region (Ref = South East) | East Midlands | 0.983 (0.795, 1.215) | 0.873 |
| Region (Ref = South East) | East of England | 0.874 (0.708, 1.080) | 0.212 |
| Region (Ref = South East) | Greater London | 1.435 (1.207, 1.707) | 0.000 |
| Region (Ref = South East) | North East | 1.010 (0.784, 1.300) | 0.941 |
| Region (Ref = South East) | North West | 0.999 (0.830, 1.203) | 0.993 |
| Region (Ref = South East) | Northern Ireland | 1.092 (0.808, 1.476) | 0.568 |
| Region (Ref = South East) | Scotland | 0.956 (0.770, 1.187) | 0.684 |
| Region (Ref = South East) | South West | 0.894 (0.719, 1.113) | 0.317 |
| Region (Ref = South East) | Wales | 0.783 (0.594, 1.032) | 0.082 |
| Region (Ref = South East) | West Midlands | 1.209 (1.003, 1.457) | 0.046 |
| Region (Ref = South East) | Yorkshire and the Humber | 0.927 (0.758, 1.133) | 0.459 |
| Income (Ref = £25,000-£34,999) | £15,000 to £24,999 | 1.024 (0.875, 1.197) | 0.771 |
| Income (Ref = £25,000-£34,999) | 35,000 to £44,000 | 0.855 (0.729, 1.002) | 0.053 |
| Income (Ref = £25,000-£34,999) | £45,000 to £54,999 | 0.827 (0.699, 0.979) | 0.027 |
| Income (Ref = £25,000-£34,999) | £55,000 to £64,999 | 0.830 (0.688, 1.001) | 0.051 |
| Income (Ref = £25,000-£34,999) | £65,000 to £99,999 | 0.749 (0.626, 0.896) | 0.002 |
| Income (Ref = £25,000-£34,999) | Do not wish to answer | 0.858 (0.665, 1.105) | 0.235 |
| Income (Ref = £25,000-£34,999) | Over £100,000 | 0.837 (0.649, 1.079) | 0.170 |
| Income (Ref = £25,000-£34,999) | Under £15,000 | 1.286 (1.076, 1.537) | 0.006 |
| Education (Ref = Level 1-3) | Do not wish to answer | 0.834 (0.453, 1.536) | 0.560 |
| Education (Ref = Level 1-3) | Level 4 | 0.732 (0.398, 1.349) | 0.318 |
| Education (Ref = Level 1-3) | None/ other | 0.928 (0.499, 1.726) | 0.814 |
| Ethnicity (Ref = White) | Asian/ Asian British | 0.828 (0.656, 1.044) | 0.111 |
| Ethnicity (Ref = White) | Black/ Black British | 1.285 (1.026, 1.609) | 0.029 |
| Ethnicity (Ref = White) | Do not wish to answer | 1.085 (0.586, 2.010) | 0.796 |
| Ethnicity (Ref = White) | Mixed | 1.390 (1.086, 1.780) | 0.009 |
| Ethnicity (Ref = White) | Other | 1.350 (1.105, 1.650) | 0.003 |
| Religion (Ref = Christian) | Atheist/ agnostic | 0.498 (0.406, 0.612) | 0.000 |
| Religion (Ref = Christian) | Do not wish to answer | 1.072 (0.804, 1.430) | 0.634 |
| Religion (Ref = Christian) | Hindu | 1.774 (1.061, 2.966) | 0.029 |
| Religion (Ref = Christian) | Muslim | 1.516 (1.106, 2.078) | 0.010 |
| Religion (Ref = Christian) | Other | 0.844 (0.655, 1.087) | 0.188 |
| First Language (Ref = English) | Do not wish to answer | 2.655 (1.287, 5.477) | 0.008 |
| First Language (Ref =   English) | Other | 1.358 (1.125, 1.639) | 0.001 |
| Interaction: Year*Age | 2021: 18-24yrs | 0.737 (0.510, 1.064) | 0.103 |
| Interaction: Year*Age | 2021: 25-34yrs | 0.965 (0.732, 1.271) | 0.799 |
| Interaction: Year*Age | 2021: 45-54yrs | 1.445 (1.031, 2.026) | 0.032 |
| Interaction: Year*Age | 2021: 55+yrs | 0.769 (0.440, 1.344) | 0.357 |
| Interaction: Year*Age | 2022: 18-24yrs | 0.358 (0.155, 0.831) | 0.017 |
| Interaction: Year*Age | 2022: 25-34yrs | 0.767 (0.593, 0.994) | 0.045 |
| Interaction: Year*Age | 2022: 45-54yrs | 1.060 (0.764, 1.469) | 0.728 |
| Interaction: Year*Age | 2022: 55+yrs | 0.808 (0.411, 1.585) | 0.535 |
| Interaction: Year*Sex | 2021: Female | 1.090 (0.868, 1.370) | 0.459 |
| Interaction: Year*Sex | 2022: Female | 1.841 (1.465, 2.312) | 0.000 |
| Interaction: Year*Religion | 2021: Atheist/ agnostic | 1.254 (0.929, 1.694) | 0.139 |
| Interaction: Year*Religion | 2022: Atheist/ agnostic | 1.679 (1.268, 2.223) | 0.000 |
| Interaction: Year*Religion | 2021: Do not wish to answer | 1.136 (0.750, 1.722) | 0.547 |
| Interaction: Year*Religion | 2022: Do not wish to answer | 0.814 (0.535, 1.238) | 0.336 |
| Interaction: Year*Religion | 2021: Hindu | 0.630 (0.315, 1.261) | 0.192 |
| Interaction: Year*Religion | 2022: Hindu | 0.388 (0.172, 0.873) | 0.022 |
| Interaction: Year*Religion | 2021: Muslim | 0.900 (0.599, 1.351) | 0.610 |
| Interaction: Year*Religion | 2022: Muslim | 0.930 (0.599, 1.443) | 0.745 |
| Interaction: Year*Religion | 2021: Other | 1.229 (0.848, 1.783) | 0.276 |
| Interaction: Year*Religion | 2022: Other | 1.082 (0.748, 1.565) | 0.675 |

**Table 12: results of ordinal logistic regression predicting change in attitudes towards giving their children routine vaccines**

| **Variable group** | **Variable** | **OR (95% CI)** | **p** |
| --- | --- | --- | --- |
| Parental COVID-19 vaccine status (Ref = 3 doses) | 1 dose | 1.697 (1.046, 2.754) | 0.032 |
| Parental COVID-19 vaccine status (Ref = 3 doses) | 2 doses | 1.204 (0.966, 1.501) | 0.099 |
| Parental COVID-19 vaccine status (Ref = 3 doses) | Not vaccinated | 2.705 (2.022, 3.618) | 0.000 |
| Age group (Ref = 35-44yrs) | 18-24yrs | 0.093 (0.041, 0.210) | 0.000 |
| Age group (Ref = 35-44yrs) | 25-34yrs | 0.655 (0.529, 0.812) | 0.000 |
| Age group (Ref = 35-44yrs) | 45-54yrs | 0.848 (0.675, 1.066) | 0.157 |
| Age group (Ref = 35-44yrs) | 55+yrs | 0.934 (0.560, 1.559) | 0.794 |
| Sex (Ref = Male) | Female | 1.350 (1.096, 1.662) | 0.005 |
| Income (Ref = £25,000 - £34,999) | £15,000 to £24,999 | 0.907 (0.654, 1.258) | 0.558 |
| Income (Ref = £25,000 - £34,999) | 35,000 to £44,000 | 1.137 (0.835, 1.548) | 0.415 |
| Income (Ref = £25,000 - £34,999) | £45,000 to £54,999 | 1.432 (1.033, 1.985) | 0.031 |
| Income (Ref = £25,000 - £34,999) | £55,000 to £64,999 | 1.373 (0.969, 1.945) | 0.075 |
| Income (Ref = £25,000 - £34,999) | £65,000 to £99,999 | 1.370 (0.986, 1.903) | 0.061 |
| Income (Ref = £25,000 - £34,999) | Do not wish to answer | 1.442 (0.902, 2.304) | 0.126 |
| Income (Ref = £25,000 - £34,999) | Over £100,000 | 1.232 (0.767, 1.981) | 0.388 |
| Income (Ref = £25,000 - £34,999) | Under £15,000 | 0.684 (0.461, 1.017) | 0.060 |
| Region (Ref = South East) | East Midlands | 0.920 (0.623, 1.358) | 0.675 |
| Region (Ref = South East) | East of England | 0.938 (0.638, 1.380) | 0.745 |
| Region (Ref = South East) | Greater London | 0.526 (0.369, 0.750) | 0.000 |
| Region (Ref = South East) | North East | 0.918 (0.576, 1.465) | 0.720 |
| Region (Ref = South East) | North West | 0.847 (0.600, 1.196) | 0.346 |
| Region (Ref = South East) | Northern Ireland | 1.549 (0.798, 3.007) | 0.196 |
| Region (Ref = South East) | Scotland | 1.095 (0.737, 1.629) | 0.653 |
| Region (Ref = South East) | South West | 0.942 (0.635, 1.395) | 0.764 |
| Region (Ref = South East) | Wales | 0.885 (0.548, 1.429) | 0.617 |
| Region (Ref = South East) | West Midlands | 0.993 (0.688, 1.434) | 0.972 |
| Region (Ref = South East) | Yorkshire and the Humber | 1.001 (0.688, 1.456) | 0.997 |
| Working status (Ref = full-time) | Do not wish to answer | 0.191 (0.039, 0.950) | 0.043 |
| Working status (Ref = full-time) | Looking after the home | 1.153 (0.834, 1.594) | 0.388 |
| Working status (Ref = full-time) | Part-time | 1.526 (1.199, 1.944) | 0.001 |
| Working status (Ref = full-time) | Retired/ disabled | 1.526 (0.944, 2.465) | 0.084 |
| Working status (Ref = full-time) | Student | 1.415 (0.525, 3.810) | 0.492 |
| Working status (Ref = full-time) | Unemployed | 1.408 (0.743, 2.667) | 0.294 |
| Ethnicity (Ref = White) | Asian/ Asian British | 1.026 (0.640, 1.644) | 0.915 |
| Ethnicity (Ref = White) | Black/ Black British | 0.616 (0.395, 0.962) | 0.033 |
| Ethnicity (Ref = White) | Do not wish to answer | 0.902 (0.211, 3.866) | 0.890 |
| Ethnicity (Ref = White) | Mixed | 0.683 (0.389, 1.200) | 0.185 |
| Ethnicity (Ref = White) | Other | 1.235 (0.788, 1.937) | 0.357 |
| Education (Ref = Level 1-3) | Do not wish to answer | 1.713 (0.459, 6.396) | 0.423 |
| Education (Ref = Level 1-3) | Level 4 | 0.898 (0.734, 1.097) | 0.292 |
| Education (Ref = Level 1-3) | None/ other | 1.050 (0.768, 1.436) | 0.760 |
| Religion (Ref = Christian) | Atheist/ agnostic | 1.586 (1.278, 1.969) | 0.000 |
| Religion (Ref = Christian) | Do not wish to answer | 1.204 (0.849, 1.707) | 0.298 |
| Religion (Ref = Christian) | Hindu | 0.268 (0.140, 0.512) | 0.000 |
| Religion (Ref = Christian) | Muslim | 0.645 (0.399, 1.044) | 0.074 |
| Religion (Ref = Christian) | Other | 1.552 (1.138, 2.119) | 0.006 |
| First Language (Ref = English) | Do not wish to answer | 0.734 (0.116, 4.650) | 0.743 |
| First Language (Ref = English) | Other | 0.687 (0.448, 1.055) | 0.086 |

**Questionnaire (2020)**

**Your views on the coronavirus/COVID-19 pandemic**  

You are invited to participate in this survey being conducted by researchers at Imperial College London to investigate the UK’s views towards the coronavirus/COVID-19 pandemic.  Your participation is **entirely voluntary**, and all your responses are **completely anonymous**. We are not collecting any information which can be used to identify you, such as your name, e-mail address, or home address.

**The survey will take between 5 and 10 minutes to complete.**

If you wish, you can exit the survey at any time, and we will not record your responses. Once you have submitted your responses, you will not be able to withdraw from the survey. Your responses are highly valuable to us and will be used to understand how adults in the UK feel about the coronavirus/COVID-19 pandemic. If you have any questions about this study, please contact us at.

If you wish to participate please click the "Continue" button below. By clicking "Continue" you are indicating that you agree to participate. You must be at least 18 years old and reside in the UK.

DEMAGE How old are you?

▼ 18 (1) ... 100 (83)

DEMREG Which UK region do you live in?

▼ East Midlands (1) ... Yorkshire and The Humber (11)

DEM We would like to ask you some questions about yourself.

DEMSEX I am

- Male (1)
- Female (2)
- Other (3)

DEMEDU What is the highest level of education you have completed? (Select the response that best applies)

- No academic qualifications (1)
- 0-4 GCSE, O-levels, or equivalents (2)
- 5+ GCSE, O-levels, 1 A level, or equivalents (3)
- Apprenticeship (4)
- 2+ A levels or equivalents (5)
- Undergraduate or postgraduate degree, or other professional qualification (6)
- Other (e.g. vocational, foreign qualifications) (7)
- Do not know (8)
- Do not wish to answer (9)

DEMWRK Which of the following best describes your work status 6 months ago?

- Working full-time (including self-employed) (1)
- Working part-time (including self-employed) (2)
- Unemployed (3)
- Student (4)
- Looking after the home (5)
- Retired (6)
- Unable to work (e.g. short- or long-term disability) (9)
- Do not wish to answer (8)

DEMREL Do you consider yourself

- Christian (1)
- Hindu (2)
- Muslim (3)
- Jewish (4)
- Buddhist (5)
- Atheist or agnostic (6)
- Other (7)
- Do not wish to answer (8)

DEMETH Which best describes your ethnicity? (select the response that best applies)

- White: English/Welsh/Scottish/Northern Irish/British (1)
- White: Irish (2)
- White: Other white background (3)
- White and Black Caribbean (4)
- White and Black African (5)
- White and Asian or White and Asian British (6)
- Black, African, Caribbean or Black British (12)
- Asian or Asian British: Indian (7)
- Asian or Asian British: Pakistani (8)
- Asian or Asian British: Chinese (10)
- Asian or Asian British: Other (11)
- Other (13)
- Do not wish to answer (14)

DEMLAN What is your first language?

- English or Welsh (1)
- Polish (2)
- Punjabi (3)
- Urdu (4)
- Bengali (5)
- Other (6)
- Do not wish to answer (7)

DEMINC What is your total household income in GBP (£) from all sources before tax?

- Under £15,000 (1)
- £15,000 to £24,999 (2)
- £25,000 to £34,999 (3)
- £35,000 to £44,999 (4)
- £45,000 to £54,999 (5)
- £55,000 to £64,999 (6)
- £65,000 to £99,999 (7)
- Over £100,000 (8)
- Do not wish to answer (9)

End of Bloc

VAC_DEC Are you currently responsible for decisions relating to the vaccination of children?

- Yes (1)
- No (2)

VAC_CHI_REFUSE Are there any immunisations you have not, or would not, allow children to have?

- Yes (1)
- No (2)
- Do not know (7)

VAC_CHI_WHICH Which vaccines, if any, do you have concerns about? (select all that apply)

- The measles, mumps, and rubella combined vaccine (MMR) (1)
- The seasonal influenza vaccine (2)
- The human papilloma virus vaccine (HPV) (4)
- Other, please specify (5) ________________________________________________
- ⊗None of the above (8)

End of Block

**Questionnaire (2021)**

**Your views on the coronavirus/COVID-19 pandemic**  

You are invited to participate in this survey being conducted by researchers at the London School of Hygiene and Tropical Medicine (LSHTM) to investigate the UK’s views towards the coronavirus/COVID-19 pandemic.  
Your participation is **entirely voluntary**, and all your responses are **completely anonymous**. We are not collecting any information which can be used to identify you, such as your name, e-mail address, or home address.

**The survey should take between 7 and 10 minutes to complete.** 

If you wish, you can exit the survey at any time, and we will not record your responses. Once you have submitted your responses, you will not be able to withdraw from the survey.    **Your responses are highly valuable to us and will be used to understand how adults in the UK feel about the coronavirus/COVID-19 pandemic.** All your anonymous responses will be made available for academic use to better understand the pandemic in the UK and its impact on beliefs and behaviours.

If you wish to participate please click the next button “->” below. By clicking next you are indicating that you agree to participate and are happy to for your anonymous data to be shared with other academic researchers. You must be at least 18 years old and reside in the UK.

DEM We will begin by asking you some questions about yourself

DEMAGENUM How old are you?

▼ 18 (1) ... 100 (83)

DEMREG Which UK region do you live in?

▼ East Midlands (1) ... Other (for example, Jersey, Guernsey, Isle of Man) (13)

DEMSEX I am

- Male (1)
- Female (2)
- Other (3)

DEMEDU What is the highest level of education you have completed? (Select the response that best applies)

- No academic qualifications (1)
- 0-4 GCSE, O-levels, or equivalents (2)
- 5+ GCSE, O-levels, 1 A level, or equivalents (3)
- Apprenticeship (4)
- 2+ A levels or equivalents (5)
- Undergraduate or postgraduate degree, or other professional qualification (6)
- Other (e.g. vocational, foreign qualifications) (7)
- Do not know (8)
- Do not wish to answer (9)

DEMWRK Which of the following best describes your work status 6 months ago?

- Working full-time (including self-employed) (1)
- Working part-time (including self-employed) (2)
- Unemployed (3)
- Student (4)
- Looking after the home (5)
- Retired (6)
- Unable to work (including, for example, a short- or long-term disability) (9)
- Do not wish to answer (8)

DEMREL Do you consider yourself

- Christian (1)
- Hindu (2)
- Muslim (3)
- Jewish (4)
- Buddhist (5)
- Atheist or agnostic (6)
- Other (7)
- Do not wish to answer (8)

DEMETH Which best describes your ethnicity (select the response that best applies)

- White: English/Welsh/Scottish/Northern Irish/British (1)
- White: Irish (2)
- White: Other white background (3)
- White and Black Caribbean (4)
- White and Black African (5)
- White and Asian or White and Asian British (6)
- Black, African, Caribbean or Black British (12)
- Asian or Asian British: Indian (7)
- Asian or Asian British: Pakistani (8)
- Asian or Asian British: Chinese (10)
- Asian or Asian British: Other (11)
- Gypsy or Irish traveller (16)
- Other (13)
- Do not wish to answer (14)
- Roma (15)

DEMLAN What is your main language

- English or Welsh (1)
- Polish (2)
- Punjabi (3)
- Urdu (4)
- Bengali (5)
- Other (6)
- Do not wish to answer (7)

DEMINC What is your total household income in GBP (£) from all sources before tax?

- Under £15,000 (1)
- £15,000 to £24,999 (2)
- £25,000 to £34,999 (3)
- £35,000 to £44,999 (4)
- £45,000 to £54,999 (5)
- £55,000 to £64,999 (6)
- £65,000 to £99,999 (7)
- Over £100,000 (8)
- Do not wish to answer (9)

Start of Block

VAC_DEC Are you **currently** responsible for decisions relating to the vaccination of children?

- Yes (1)
- No (2)

VAC_CHI_REFUSE Are there any immunisations you have not, or would not allow children to have?

- Yes (1)
- No (2)
- Do not know (3)

VAC_CHI_WHICH Which vaccines, if any, do you have concerns about (select all that apply)

- The measles, mumps, and rubella combined vaccine (MMR) (1)
- The seasonal influenza vaccine (2)
- The human papilloma virus vaccine (HPV) (3)
- The coronavirus (COVID-19) vaccines (4)
- Other, please specify (5) ________________________________________________
- ⊗None of the above (6)

**Questionnaire (2022)**

**Your views on the coronavirus/COVID-19 pandemic**  

You are invited to participate in this survey being conducted by researchers at the London School of Hygiene and Tropical Medicine (LSHTM) to investigate the UK’s views towards the coronavirus/COVID-19 pandemic.  

Your participation is **entirely voluntary**, and all your responses are **completely anonymous**. We are not collecting any information which can be used to identify you, such as your name, e-mail address, or home address. 

**The survey should take no longer than 5 minutes to complete.**

If you wish, you can exit the survey at any time, and we will not record your responses. Once you have submitted your responses, you will not be able to withdraw from the survey.    **Your responses are highly valuable to us and will be used to understand how adults in the UK feel about the coronavirus/COVID-19 pandemic.**    All your anonymous responses will be made available for academic use to better understand the pandemic in the UK, but as a reminder, your responses will **always stay completely anonymous**.

If you wish to participate please click the next button “->” below. By clicking next you are indicating that you agree to participate and are happy to for your anonymous data to be shared with other academic researchers. You are also acknowledging that you are at least 18 years old and reside in the UK.

VAC_DEC Are you currently responsible for decisions relating to the vaccination of children?

- Yes (1)
- No (2)

SOCDEM We will begin by asking you some questions about yourself

AGE How old are you?

▼ 18 (1) ... 100 (83)

REG Which UK region do you live in?

▼ East Midlands (1) ... Other (for example, Jersey, Guernsey, Isle of Man) (13)

SEX I am

- Male (1)
- Female (2)
- Other (3)

EDU What is the highest level of education you have completed? (Select the response that best applies)

- No academic qualifications (1)
- 0-4 GCSE, O-levels, or equivalents (2)
- 5+ GCSE, O-levels, 1 A level, or equivalents (3)
- Apprenticeship (4)
- 2+ A levels or equivalents (5)
- Undergraduate or postgraduate degree, or other professional qualification (6)
- Other (e.g. vocational, foreign qualifications) (7)
- Do not know (8)
- Do not wish to answer (9)

EMP Which of the following best describes your work status 6 months ago?

- Working full-time (including self-employed) (1)
- Working part-time (including self-employed) (2)
- Unemployed (3)
- Student (4)
- Looking after the home (5)
- Retired (6)
- Unable to work (including, for example, a short- or long-term disability) (9)
- Do not wish to answer (8)

REL Do you consider yourself

- Christian (1)
- Hindu (2)
- Muslim (3)
- Jewish (4)
- Buddhist (5)
- Atheist or agnostic (6)
- Other (7)
- Do not wish to answer (8)

ETH Which best describes your ethnicity (select the response that best applies)

- White: English/Welsh/Scottish/Northern Irish/British (1)
- White: Irish (2)
- White: Other white background (3)
- White and Black Caribbean (4)
- White and Black African (5)
- White and Asian or White and Asian British (6)
- Black, African, Caribbean or Black British (12)
- Asian or Asian British: Indian (7)
- Asian or Asian British: Pakistani (8)
- Asian or Asian British: Chinese (10)
- Asian or Asian British: Other (11)
- Gypsy or Irish traveller (16)
- Roma (15)
- Other (13)
- Do not wish to answer (14)

LAN What is your main language

- English or Welsh (1)
- Polish (2)
- Punjabi (3)
- Urdu (4)
- Bengali (5)
- Other (6)
- Do not wish to answer (7)

INC What is your total **household** income in GBP (£) from all sources before tax?

- Under £15,000 (1)
- £15,000 to £24,999 (2)
- £25,000 to £34,999 (3)
- £35,000 to £44,999 (4)
- £45,000 to £54,999 (5)
- £55,000 to £64,999 (6)
- £65,000 to £99,999 (7)
- Over £100,000 (8)
- Do not wish to answer (9)

Start of Block: Childhood immunisations: non-COVID (1)

VAC_CHI_REFUSE Are there any immunisations you have not, or would not, allow children to have?

- Yes (1)
- No (2)
- Do not know (4)

VAC_CHI_WHICH Which vaccines, if any, do you have concerns about? (select all that apply)

- The measles, mumps, and rubella combined vaccine (MMR) (1)
- The seasonal influenza vaccine (2)
- The coronavirus (COVID-19) vaccines (6)
- The human papilloma virus vaccine (HPV) (3)
- Other, please specifiy (4) ________________________________________________
- ⊗None of the above (5)

CHILD_CONF Since the beginning of the COVID-19 pandemic, has your attitude towards giving your child/children routine vaccines changed? (The Covid-19 vaccine is not considered a routine vaccination.)

- Yes (1)
- No (2)
- Do not know (3)

CHILD_CONF_HOW How have your attitudes towards other vaccines for your child/children changed?

- I feel much more positive about giving my child vaccines (1)
- I feel slightly more positive about giving my child vaccines (2)
- I feel slightly less positive about giving my child vaccines (3)
- I feel much less positive about giving my child vaccines (4)
